# Shifting PrEP Initiation from Practitioner to Nurse: a clinic-level multi-modal intervention protocol to increase PrEP uptake among women at increased risk of HIV acquisition in a U.S. OB/GYN clinical setting

**DOI:** 10.1101/2022.07.14.22277560

**Authors:** Erin L. Gingher, Timothee F. Fruhauf, Saumya S. Sao, Runzhi Wang, Jenell S. Coleman

**Affiliations:** Johns Hopkins School of Medicine

**Keywords:** pre-exposure prophylaxis (PrEP), HIV prevention, Ending the HIV Epidemic (EHE), nurse, women

## Abstract

Women comprise 20% of new HIV diagnoses in the U.S. with 86% attributed to heterosexual contact, but HIV pre-exposure prophylaxis (PrEP) uptake is low. OB/GYN clinics are ideal settings to increase HIV prevention counseling for cisgender women, as patients are more likely to discuss their sexual behavior, undergo sexually transmitted infection screening, and receive risk reduction counseling. Our study will assess the feasibility, acceptability, and effectiveness of a registered nurse (RN)-led PrEP project in OB/GYN clinics. Microlearning and Plan-Do-Study-Act cycles will be performed, followed by a randomized controlled trial (RCT). A total of cisgender women determined to be at-risk for HIV will be randomized to standard of care with electronic medical record enhancements (e-SOC) or e-SOC with contact and PrEP counselling by an RN, who will be able to prescribe PrEP under protocol guidance. By shifting HIV PrEP counselling to a nurse, clinics may be able to increase PrEP awareness and uptake.

Registered with clinicaltrials.gov: NCT05095818

## Background

HIV remains a significant public health threat in the United States (U.S.). Despite increased testing, prevention, and treatment services, the number of Americans newly infected with HIV remains stable, with approximately 38,000 diagnosed each year (HIV.gov, 2021). Women^1^ represent nearly one in five of these new diagnoses, and Black women are disproportionately affected (60%) (Carley et al., 2019).

There are currently several effective HIV prevention strategies, including needle exchange programs, consistent condom use, and prevention medications, such as pre-exposure prophylaxis (PrEP) and post-exposure prophylaxis (PEP). In 2010, the iPrEx study reported reduced risk of HIV acquisition with oral emtricitabine/tenofovir disoproxil fumarate (FTC/TDF) among HIV-seronegative men and transgender women who have sex with men (Grant et al., 2010). Shortly thereafter, the U.S. Food and Drug Administration (FDA) approved once daily oral FTC/TDF as PrEP for HIV prevention in 2012 (FDA, n.d.). With high daily adherence, PrEP is over 90% effective at preventing HIV acquisition. The U.S. Public Health Service released a 2021 clinical practice guideline update for PrEP that recommends informing all sexually active adults and adolescents about PrEP. Despite the proven effectiveness of PrEP, utilization remains low among heterosexual women. In 2018, only 7% of women in the U.S. who could benefit from PrEP were prescribed it (CDC, 2021).

Multilevel barriers are associated with poor PrEP uptake among women. At the patient level, barriers include minimal knowledge of PrEP and underestimation of HIV acquisition risk (Aaron et al., 2018; Auerbach et al., 2015; Johnson et al., 2020; Ojikutu & Mayer, 2019). In a study that surveyed urban obstetrics & gynecology (OB/GYN) clinics in Louisiana, the majority of patients perceived their risk of HIV acquisition to be low, despite known risk factors, such as inconsistent condom use with sexual activity in a region of high HIV prevalence (Carley et al., 2019). Another study assessing the sexual behavior of pregnant women in Washington, D.C. found no association between the number of behavioral risk factors and perceived risk of HIV acquisition (Scott et al., 2021).

Once informed about PrEP, women express concern over potential side effects, dislike of swallowing pills, and costs. The burden of frequent healthcare appointments and stigma associated with taking a medication used by people living with HIV may also impede adoption of PrEP (Collier et al., 2017; Hirschhorn et al., 2020). Interpersonal relationships, such as the influence of sexual partners and practitioners, have also been identified as potential individual-level barriers to PrEP (Ogunbajo et al., 2021)

Furthermore, barriers at the practitioner-level exist, including discomfort in taking a sexual history, decreasing clinic visit length, competing clinical demands, and lack of knowledge (O’Byrne et al., 2019). However, even when practitioners are knowledgeable about PrEP, uptake is low. For example, in our own clinics, we found that approximately 10% of pregnant patients were eligible for PrEP, but none were given prescriptions (Fruhauf & Coleman, 2017). With an understanding of the PrEP barriers for heterosexual women and the federal government’s *Ending the HIV Epidemic* (EHE) Initiative, which aims to reduce the number of new HIV infections by at least 90 percent by 2030, we sought to increase PrEP awareness and uptake in our outpatient OB/GYN clinics.

This manuscript describes our clinic-level multi-modal intervention protocol to increase HIV prevention services within OB/GYN clinical settings. We plan to conduct microlearning sessions, Plan-Do-Study-Act cycles, and a pragmatic randomized controlled trial (RCT).

## Methods

### Setting

This project will be launched in three OB/GYN outpatient clinics of a single academic medical system in Baltimore, Maryland, which has a Black majority population and a poverty rate of over 20% (U.S. Census Bureau, n.d.). In 2018, Baltimore City had an estimated HIV incidence rate of 42.1 per 100,000 population (CDC, 2020). It is one of the 57 jurisdictions targeted in the EHE Initiative (HIV.gov, 2020). The clinics offer general obstetric and gynecologic care, with some providing specialized care for high-risk pregnancies. We selected an outpatient OB/GYN setting because it is where women routinely receive STI counseling, testing, and treatment. Data has shown that STIs increase the risk of HIV acquisition and transmission (Ward & Rönn, 2010). Furthermore, HIV incidence among pregnant and postpartum women is 2-4 times higher than the rates outside of pregnancy, likely due to behavioral and biological changes during this time period (Mugo et al., 2011; Thomson et al., 2018). Therefore, regularly scheduled obstetric and annual gynecologic visits are opportunities for healthcare practitioners to discuss sexual health and HIV-risk behaviors with patients.

### Patients

Patients will be included in the project if they are assigned female at birth, not living with HIV, and are receiving OB/GYN care in our preselected clinics. Obstetric patients will be identified through an existing non-validated STI/HIV risk assessment tool embedded into our electronic medical record (EMR) (**Table 1**), which is completed by an RN during a patient’s initial prenatal care visit. This tool was developed over a decade ago as part of general behavioral risk assessment for obstetrics patients, and not specifically for PrEP eligibility.

**Table 1:** Obstetric HIV/STI Risk Tool>.

| Risk factor |  | Response options |  |
| --- | --- | --- | --- |
| Inject illicit drugs | **Yes, in the past 6 months** | **Yes, more than 6 months ago** | **Never** |
| Sex with partner who injects illicit drugs | **Yes, in the past 6 months** | **Yes, more than 6 months ago** | **Never** |
| Any past STIs | **Yes, in the past 6 months** | **Yes, more than 6 months ago** | **Never** |
| Sex with a partner who has sex with both men and women | **Yes, in the past 6 months** | **Yes, more than 6 months ago** | **Never** |
| Sex for money, drugs, other payment | **Yes, in the past 6 months** | **Yes, more than 6 months ago** | **Never** |
| Sex with a partner currently infected with HIV | **Yes, in the past 6 months** | **Yes, more than 6 months ago** | **Never** |
| Sex with more than one partner | **Yes, in the past 6 months** | **Yes, more than 6 months ago** | **Never** |

Eligible gynecologic patients will be identified by a positive STI diagnosis within the past 6 months, or if seeking STI testing and have a history of an STI (either self-reported or resulted in the EMR). Because PrEP eligibility criteria (**Table 2**) differ by guidelines and expert opinion, we selected broad criteria given the high HIV prevalence in our area (U.S. Public Health Service, 2018; The American College of Obstetricians and Gynecologists [ACOG], 2014). Before PrEP is prescribed, PrEP contraindications will be followed, including: symptoms or clinical signs consistent with acute HIV infection, unknown HIV infection status, allergies to the active substances or any excipients of PrEP, estimated creatinine clearance of <60 mL/minute, active Hepatitis B infection, or are participating in other clinical studies related to HIV and/or antiretroviral therapy.

**Table 2:** PrEP Eligibility Criteria by Organization>.

| Risk Factors | ACOG | CDC | USPSTF | WHO* |
| --- | --- | --- | --- | --- |
| Sex in high HIV-prevalence area or social network (plus at least one other risk factor) | ✓ |  |  |  |
| Inconsistent or no condom use | ✓ | ✓ | ✓<br>(inconsistent condom use with a partner whose HIV status is unknown and who is at high risk, such as injection drug user or man who has sex with men and women) | ✓ |
| Sexual partner with unknown HIV status | ✓<br>(who has at least one other risk factor) |  |  | ✓<br>(and partner is high risk) |
| Sexual partner living with HIV | ✓ | ✓ | ✓ | ✓ |
| STI diagnosis within past 6 months | ✓<br>(no time frame) | ✓ | ✓<br>(syphilis or gonorrhea) | ✓ |
| Injection drug use | ✓ | ✓<br>(with shared injection equipment or injection partner with HIV) | ✓<br>(with shared drug injection equipment or risk of sexual acquisition) | ✓<br>(with shared needles/syringes) |
| Recreational drug use |  |  | ✓<br>(with shared drug injection equipment or risk of sexual acquisition) | ✓<br>(sex under the influence of recreational drugs) |
| Alcohol dependence | ✓ |  |  | ✓<br>(sex under the influence of alcohol) |
| Incarceration | ✓ |  |  |  |
| Transactional sex | ✓ |  |  | ✓ |
| Ongoing IPV/GBV |  |  |  | ✓ |
| Recurrent use of PEP |  | ✓ |  | ✓ |

ACOG: The American College of Obstetricians and Gynecologists
CDC: Centers for Disease Control and Prevention
USPSTF: United States Preventive Services Task Force
WHO: World Health Organization
IPV: intimate partner violence
GBV: gender-based violence
PEP: post-exposure prophylaxis
\*According to WHO, PrEP eligibility may vary by country. HIV-negative individuals who are “determined to be at substantial risk for HIV as defined by national guidelines” are eligible for PrEP. WHO does not list specific criteria but offers an example tool for determining eligibility. These example criteria are included in the table.

### Study design

The study is designed with several, sequential methodologic components because of our organizational culture and prior data that suggest complex, multi-level barriers to PrEP uptake among women. First, we will synthesize the existing literature, engage key stakeholders, and conduct microlearning sessions about PrEP. Microlearning sessions involve the delivery of educational content that is brief, targeted, and occur within clinical sites (Buchem & Hamelmann, 2010). Microlearning sessions will be conducted among all healthcare team members in the clinic, which includes nurses, medical assistants, nurse practitioners, midwives, and physicians. Then, Plan-Do-Study-Act (PDSA) will be used. The PDSA method is a commonly used improvement process in health care settings, which utilizes small tests of change to optimize a process (Coury et al., 2017). A common misperception with PDSA methodology is that it is a standalone method (Reed & Card, 2016). Therefore, we designed multiple rigorous methodologies to include along with PDSA.

The first PDSA cycle will involve EMR changes and identification of a registered nurse (RN) who will be trained to initiate PrEP. Our prior data has shown that PrEP is rarely discussed or prescribed to patients, despite indications (Fruhauf & Coleman, 2017; Lopez et al., 2019). Proposed EMR changes will include the creation of (1) Best practice alerts (BPAs) that encourage nurses and practitioners to add HIV risk-related ICD-10 codes to the problem lists. Problem lists contain a summary snapshot of the patient’s medical conditions for use by all practitioners. A positive risk screen for prenatal care patients will alert the nurse to add “HIV risk factors complicating pregnancy” to the patient’s problem list during the initial prenatal visit. When a patient tests positive for an STI and is prescribed an antibiotic for treatment, a BPA will alert the GYN practitioner that the patient may have HIV acquisition risk factors and to add “encounter for HIV counseling” to the problem list; (2) Comprehensive and easily accessible order sets that facilitate initiation of PrEP by outlining the required laboratory tests and their timing, and the exact FTC/TDF medication instructions to avoid incorrect prescriptions; (3) Note templates that allow for consistent documentation and serve as an educational tool to ensure adequate HIV prevention counseling is complete; (4) PrEP patient educational material to include in patient after-visit summaries, which are given to patients at the conclusion of clinic appointments. Educational materials include frequently asked questions (FAQ), FTC/TDF medication facts, and PrEP use during pregnancy or breastfeeding; and (5) short animation videos that describe PrEP.

We will identify a PrEP-RN who will undergo training in HIV prevention, including the use of PrEP. Well within the scope of nursing practice, RNs can screen patients for risk factors, counsel and educate at-risk patients about effective HIV prevention strategies, and advocate for PrEP utilization. To our knowledge, this is the first U.S. RN-led PrEP program for women exclusively. One novel aspect of our program is that our PrEP-RN will be able to initiate laboratory test ordering and PrEP prescriptions, under specific protocol guidance and supervision by the clinic’s Medical Director. Another innovative aspect is that we designed our PrEP-RN program to allow for telehealth counseling sessions for initiation and follow-up sessions via telephone or video. Patients are only required to visit a brick-and-mortar location to complete laboratory tests, which can take place at a location most convenient for them.

Other models for RN-led PrEP programs have been developed in Canada, but none focus exclusively on women, take place in an OB/GYN outpatient setting where women prefer to receive information about HIV prevention, or allow telehealth (Auerbach et al., 2015; Hirschhorn et al., 2020; Sharma et al., 2018). As O’Byrne highlights in their PrEP-RN model, the numerous appointments involved with the first year of PrEP use has a high potential for missed appointments, which may be remedied by the use of telehealth (O’Byrne et al., 2019).

The second PDSA cycle will incorporate feedback from the first cycle and then deploy the PrEP-RN intervention at the main outpatient clinic site. Subsequent PDSA cycles will include roll-out of the project at the remaining clinical sites with protocol adjustments, as needed based on the prior PDSA cycle. To test whether an RN can effectively counsel patients about PrEP, initiate therapy, and follow patients with minimal practitioner involvement, we plan to conduct a pragmatic randomized controlled trial (RCT). A pragmatic (effectiveness) trial was selected to generate evidence that would be applicable in a real-world clinical practice setting and increase external validity. The control arm will include an enhanced standard of care (e-SOC), which is standard of care plus the EMR changes. The intervention arm will include the PrEP-RN plus e-SOC.

Patients randomized to the intervention arm will first be contacted via an electronic patient portal integrated with the EMR that allows patients access to their medical records and a messaging system to communicate with practitioners. While most of our patients utilize this patient portal, if a patient does not have access, this step will be skipped.

Follow-up telephone calls will then be made to counsel patients on HIV risk factors in general, their HIV risk factors in particular, and prevention methods, inclusive of PrEP. PrEP, including same-day PrEP, will be offered. If the required laboratory tests have not been obtained in the past month, the PrEP-RN will place and order the laboratory tests. The PrEP-RN will utilize the PrEP order set in the EMR to order labs, prescribe medication electronically to the patient’s pharmacy, provide patient education, and schedule follow-up appointments. All orders and prescriptions are sent to the clinic’s Medical Director for authorization. Standardized smartphrases for clinical notes will also be utilized to record the interaction by the PrEP-RN with the study participants.

If a patient declines PrEP, no further attempts are made by the PrEP-RN to counsel the patient. Additional educational material may be sent via the electronic patient portal as desired by the patient. The patient is encouraged to contact the PrEP-RN, via telephone or the electronic patient portal, if she has any future questions or decides to use PrEP.

A Manual of Procedures (**Appendix A**) was created for the PrEP-RN to carry out each step of PrEP initiation and follow up. The manual outlines how to respond to abnormal tests, provide prescription coverage assistance, and coordinate care with other specialties as needed to address test results. The PrEP-RN will be responsible for follow-up visits and will contact the patients one-two weeks after medication initiation. At this time, the PrEP-RN will review the importance of daily adherence to PrEP and assess for any side effects.

### Randomization and blinding

Stratified block randomization will be used to allocate eligible patients into the intervention and control arms with a 1:1 ratio. Patients will be classified by patient type (obstetric or gynecologic) and clinics. To avoid imbalance between the two study arms, the same type of patients from the same site will be randomized within a block that comprise four ordered assignments. SAS will be used to generate the randomization table. EMR data abstracts will be obtained biweekly to identify PrEP eligible patients. A member of the study team will assign these PrEP eligible patients to an intervention arm based on the randomization allocation per patient type and site. Practitioners will be blinded to study assignment to prevent interference in the standard of care delivery.

### Ethics Approval and Waiver of Consent to Participate

The study received approval from the Institutional Review Board (IRB) of Johns Hopkins Medicine. Although the primary purpose of the project was to improve patient outcomes, the IRB reviewed the project as research because we included randomization plus an external funding source. However, a waiver of consent was granted because the study involves no more than minimal risk to the subjects, the waiver does not adversely affect the rights or welfare of the subjects, and the research could not be practicably completed without the waiver.

### Sample size estimate

To test the effectiveness of the PrEP-RN, we estimate that we will require 440 patients (220 women per arm). The sample size was selected to provide 80% power to detect an effect size of 15% between the control and intervention group, at an alpha level of 0.05 using two-sided test. The PrEP-RN intervention and EMR prompts will cease once the sample size has been reached.

## Outcomes

The project will be evaluated using the Reach, Effectiveness, Adoption, Implementation, and Maintenance (RE-AIM) framework. RE-AIM has five domains: Reach the target population with the intervention; Effectiveness or impact of the intervention; Adoption by target patients, staff, clinics; Implementation of the intervention, including consistency of delivery; and Maintenance of the intervention over the long term (RE-AIM, n.d.). The Reach and Effectiveness domains are individual-level factors, while the Adoption and Implementation domains are systems-level factors. The Maintenance domain includes both individual- and systems-level factors.

### Primary Outcomes

The primary outcome estimated by the RCT will be awareness of PrEP, measured by the percentage of at-risk women with EMR documentation of HIV risk prevention counseling.

### Secondary Outcomes

Secondary outcomes will include feasibility, which will be defined as reaching the target goal in at least 3 of the RE-AIM domains (**Table 3**). Other secondary outcomes include the percentage of women at risk for HIV who initiate PrEP during the study interval and the percentage of women who are using PrEP at 3-, 6-, 9-, and 12-months from initiation. PrEP initiation will be measured by the number of PrEP laboratory tests and the number of PrEP prescriptions ordered. We will also assess the utilization of EMR enhancements, measured as the percentage of patients with an HIV-risk diagnosis added to their EMR problem list. Last, we will evaluate the acceptability of the PrEP-RN intervention among our study population.

**Table 3.** Re-AIM Evaluation>.

| Domain | Target |
| --- | --- |
| **REACH** |  |
| % women with documented HIV risk counseling, including PrEP (awareness) | ≥80% |
| **EFFECTIVENESS** |  |
| % women at-risk for HIV who agree to PrEP (uptake: PrEP lab orders and prescriptions) | ≥50% |
| % women at-risk for HIV who agree to same-day PrEP | ≥30% |
| **ADOPTION** |  |
| % of nurses adding HIV risk diagnosis to problem list | ≥20 |
| **IMPLEMENTATION** |  |
| % pregnant women with completed HIV risk screen randomized | ≥90% |
| % at-risk pregnant women with diagnosis on problem list | ≥80% |
| % STI positive women randomized | ≥90% |
| % STI positive women in intervention arm contacted by PrEP nurse | ≥75% |
| **MAINTENANCE** |  |
| % women at-risk for HIV using PrEP x 3 months | ≥35% |
| % women at-risk for HIV using PrEP x 6 months | ≥25% |
| % women at-risk for HIV using PrEP x 12 months | ≥15% |

### Acceptability

Acceptability of and satisfaction with the PrEP-RN intervention will be evaluated using a brief survey of a randomly selected group of patients (n=25) who received the PrEP-RN intervention within the previous three months (**Table 4**). Items related to patient satisfaction and telehealth acceptability were identified through a review of the literature (Weaver et al., 2021; Yip et al., 2003; Gwadry-Sridhar et al., 2003).

**Table 4.**
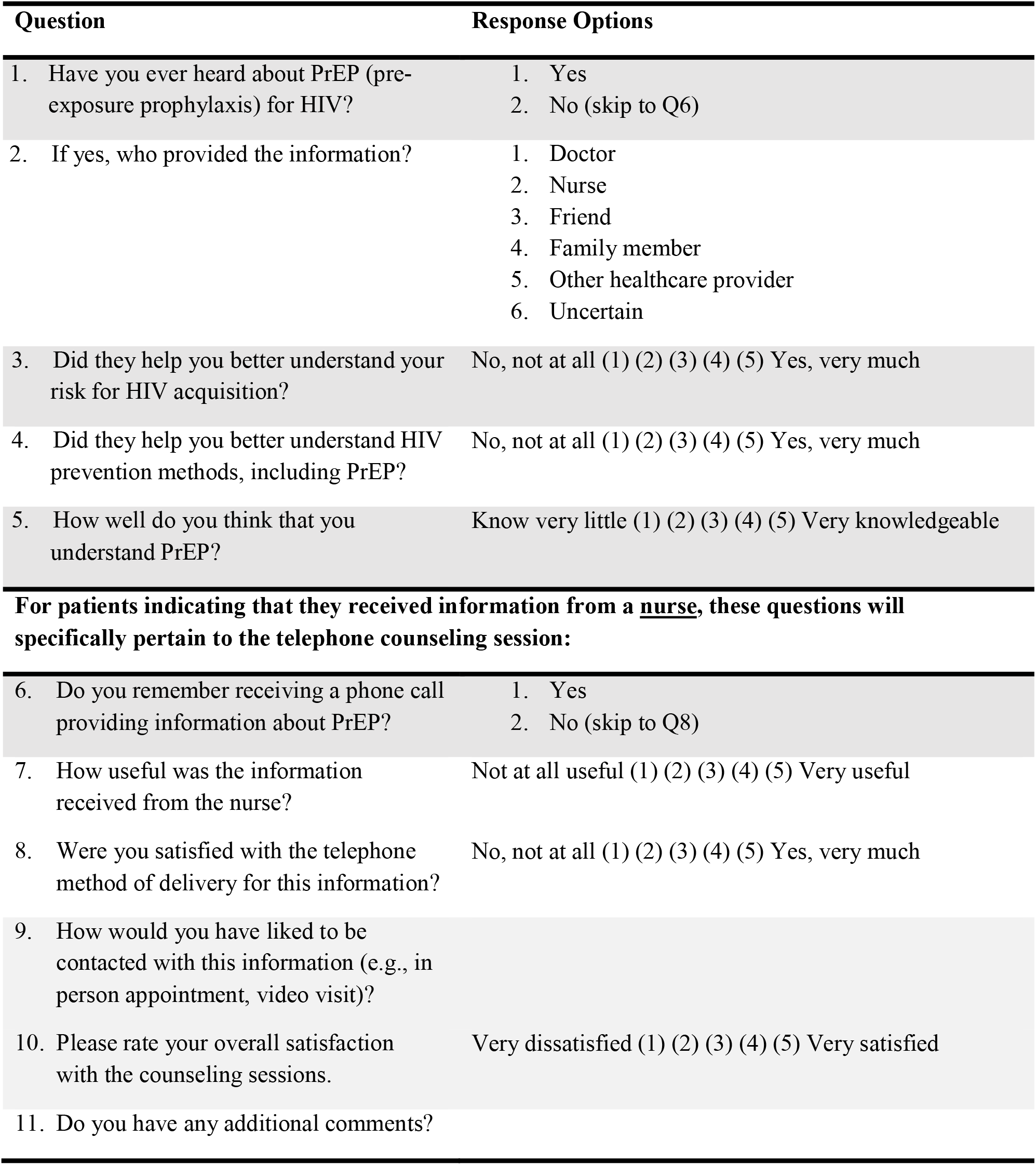
PrEP RN Patient Acceptability Questionnaire>.

### Risks and monitoring

The intervention does not pose any greater risks than routine clinical care, and no participant will receive less than e-SOC. Because the intervention is integrated at the clinic level and poses minimal risk, we do not have an independent data monitoring committee. The team will report any adverse events to the IRB. To ensure the quality of our counseling intervention, we will have weekly check-ins with the PrEP RN, research staff, and the study Principal Investigator to discuss any issues in counseling patients and incorporate patient feedback on an ongoing basis.

## Statistical Analysis

Sociodemographic characteristics of the study population will be reported using descriptive statistics. For the RCT, statistical analysis will be performed based on the intention-to-treat principle. We will compare the percentage of at-risk women with EMR documentation of HIV risk prevention counseling (i.e. primary outcome), percentage of women at risk for HIV who initiate PrEP during the study interval, and the percentage of women who are using PrEP at 3-, 6-, 9-, and 12-months from initiation between the intervention group and the control group using the chi-squared test with Fisher’s exact. Since participants in the two arms are expected to have equal baseline characteristics due to randomization, basic confounders will not be considered. However, if any baseline characteristics differ significantly between the two groups, we will use regression models to control them. Additionally, among women randomized to the PrEP RN, the reason for initiating or not initiating PrEP will be presented as a number and percentage. Also, we will plot the percentage of women who are using PrEP for 3-, 6-, 9-, and 12-months to demonstrate the trend. Marginal models will be used to identify factors associated with the trend. Lastly, to assess whether the interventions had a sustainable impact on practitioners’ behavior after withdrawing them, the primary and secondary outcomes will be compared to the year before the PrEP RN project began, using the chi-squared test.

## Discussion

Prevention is one of the four key strategies outlined by the federal government’s EHE initiative. Our novel project seeks to prevent new HIV transmissions by addressing known barriers to PrEP at the patient, practitioner, and system levels.

OB/GYN clinics were chosen for this project for several reasons. First, women have consistently expressed a preference for receiving HIV prevention services in clinics where they receive OB/GYN care, as opposed to specialty clinics that treat infectious diseases or STIs where most PrEP projects are located (Auerbach et al., 2015; Carley et al., 2019; Hirschhorn et al., 2020). Second, despite a history of mistrust in the health care system, Black and Hispanic women reported trusting their individual gynecologist (Hirschhorn et al., 2020; Kennedy et al., 2007). Third, PrEP use by women in the U.S. is largely inadequate, as the PrEP-to-need ratio for women is only 0.4 compared to 2.1 for men (Highleyman, 2018). Last, OB/GYN practitioners manage women during heightened periods of HIV acquisition risk, such as during the course of infection with an STI or during pregnancy and the postpartum period, and they routinely provide sexual and reproductive care for at-risk women (ACOG, 2018).

Clinic nurses are well suited for PrEP counseling sessions, as they are accustomed to delivering positive STI results to patients and providing risk reduction strategies. Furthermore, registered nurses represent a more abundant and less expensive resource than physicians or mid-level providers (American Association of Colleges of Nursing, 2019).

Through the role of the PrEP RN, we are utilizing telemedicine to help combat barriers to PrEP awareness and initiation. Telemedicine may improve efficiency, while also providing increased comfort and convenience for patients. Barriers to patient access, such as limited time off from work and childcare responsibilities are minimized (Hasselfeld, n.d.; Gajarawala & Pelkowski, 2021). Feasibility and efficacy, as well as patient acceptability, will be evaluated to determine the overall success of the project.

Despite the strengths, our protocol has several limitations. Our health system has EMRs, which facilitates the ease of nursing staff to place orders and prescriptions to be signed by a practitioner. Systems without EMRs may find our protocol cumbersome, however, standing orders may be considered instead. It is also plausible that changes in PrEP uptake could be due in part to practitioner behavior changes resulting from the EMR enhancements, as opposed to the PrEP-RN. Furthermore, by targeting female patients who are engaged in clinical care, we are unable to reach women at-risk for HIV who do not have access to routine care. Lastly, our urban high HIV prevalence location may limit the generalizability of this study.

Nonetheless, if this study demonstrates that a PrEP-RN can increase PrEP awareness and uptake among at-risk women, we plan to test these interventions, including the EMR enhancements in non-OB/GYN clinical settings. Our multimodal PrEP intervention could be implemented in other settings and largely improve current strategies for increasing PrEP utilization among at-risk women.

## Data Availability

This paper contains no data as it is a clinical trial protocol.

## Acknowledgements

The authors would like to acknowledge the GYN/OB clinic staff for their assistance with this study.

## Key Considerations

- There are many reasons why PrEP uptake among American women is so low, with underestimation of HIV acquisition risk a major factor.
- HIV risk factor and prevention counseling can take place via telemedicine, which has increasingly become a popular and convenient method of healthcare delivery.
- The EMR can be utilized to alert practitioners to HIV risk factors and aid in the identification of PrEP-eligible patients.

### APPENDIX A

#### PrEP RN Manual

The decision to prescribe PrEP is based on the clinical judgment of the nurse evaluating the patient. The following is a list of steps to be followed by the PrEP RN.

##### a) Determine Eligibility

Women (assigned female at birth) ≥15 years old at risk for HIV infection should be considered for PrEP and will be assessed for the option of same-day PrEP start.

- Living in (or seeking medical care) in a high HIV-prevalence area or social network
- Inconsistent or no condom use
- Specific criteria for patient type:
  - GYN patients:
    ▪ current or recent (within last six months) STI diagnosis
    ▪ lifetime history of STI and currently seeking STI testing
  - OB patients: lifetime history of at least one of the following:
    ▪ STI diagnosis
    ▪ More than one concurrent sexual partner
    ▪ Injection of illicit drugs
    ▪ Sex with someone who injects illicit drugs
    ▪ Sex with someone who has sex with both men and women
    ▪ Sex with a partner currently living with HIV
    ▪ Sex for money, drugs, or other payments
- If a patient is PEP (post-exposure prophylaxis)-eligible based on clinical guidelines, including exposure within the past 72 hours, the patient should be started on PEP and transitioned to PrEP after 1 month. Upon transition from PEP to PrEP the patient needs to be reassessed for clinical signs of HIV infection and labs for PrEP start need to be obtained.
- If a patient has any of the following conditions, she is not a candidate for PrEP:
  - unknown or known HIV infection
  - renal insufficiency (CrCl <60ml/min)
  - active Hepatitis B infection
  - signs or symptoms of acute HIV infection
  - not able to strictly adhere to the treatment or monitoring protocols
  - weight<35 kg
  - allergies to the active substances or any excipients of PrEP
  - taking PEP or other antiretroviral therapy
- If a patient meets both of the following criteria for possible acute HIV syndrome, they will not be candidates for same-day start:
  - High risk exposure to HIV within the past 6 weeks
  - Clinical signs and symptoms (currently or within the past 2 weeks) of acute HIV infection, including fever, lymphadenopathy, rash, diarrhea, oral ulcers, myalgia/arthralgia

##### b) Same-Day-PrEP-Start

- If a patient is found eligible for same-day-PrEP start, a 4^th^ generation rapid HIV test needs to be performed.
  - If negative, all PrEP initiating labs will be ordered.
  - If positive, a 4th generation HIV Antibody/Antigen and HIV viral load test should be ordered
    ▪ Patient is referred to Adult HIV Clinic immediately for ART
- Exclusion criteria for same day PrEP are:
  - Positive HIV test or clinical signs and symptoms of possible acute HIV syndrome, or possible exposure to HIV within the past 72 hours
  - No insurance. Patient will meet with case manager to discuss insurance eligibility.
  - Age >50 years AND no renal function test within the past 6 months on file
  - No working telephone number
- If patient found to eligible and interested in same-day-PrEP start, 30 days of PrEP will be ordered per protocol and pended for signature by the Medical Director.
- Patient will be counseled that there is a risk that PrEP must be stopped if labs reveal any abnormalities, such as renal impairment, in which case patient will receive a call by the nurse.
- If the patient has sexual or drug partners who are known to be HIV infected, the nurse should attempt to determine whether these partners are receiving antiretroviral therapy and assist with linkage to care for these partners if they are not in care or not receiving antiretroviral therapy.

##### c) Delayed-PrEP-Start

- If decision is made not to start PrEP on the same day routine PrEP initiation labs are ordered:
  - If the patient has symptoms consistent with acute HIV infection OR if the patient has had a risky exposure (injection drug use, unprotected intercourse) in the last three weeks, an HIV viral load test will be drawn, in addition to a 4th generation HIV Antibody/Antigen test
  - Patients without signs of acute HIV infection, a 4th generation HIV Antibody/Antigen test will be drawn
  - Patients will be advised to remain abstinent until they begin PrEP.
- The nurse will confirm that the patient’s calculated creatinine clearance is greater than or equal to 60 mL/minute (via Cockcroft-Gault formula).
  - If lower than 60 mL/minute, notify Medical Director

##### d) All Patients Starting on PrEP

- The nurse will screen for HBV infection and vaccinate against HBV if susceptible, or treat if active infection exists, regardless of the decision about prescribing PrEP.
- The nurse will screen and coordinate treatment, as needed, for STIs.
- The nurse will review and confirm potential drug interactions.
- In women of reproductive age or transmales who have sex with males, the nurse will:
  - screen for pregnancy with a urine HCG test.
  - determine if patient is planning to become pregnant or if patient is breastfeeding.

##### e) Beginning PrEP Medication Regimen

After reviewing all lab work to confirm a patient is appropriate for PrEP, the nurse will:

- Order 1 tablet of Truvada (TDF 300mg plus FTC 200mg) by mouth daily.
  - Prescribe no more than a 90-day supply, renewable only after HIV testing confirms that a patient remains HIV-uninfected.
  - Route the order to the Medical Director for signing
  - If active HBV infection is diagnosed, the nurse will notify Medical Director/ refer to Infectious Disease Specialist
- Provide risk-reduction and PrEP medication adherence counseling and condoms.
- Confirm patient’s insurance coverage. Refer uninsured patients or patients with high prescription co-pays to https://www.pleaseprepme.org/payment
- Place ICD 10 Code on problem list (Encounter for HIV counseling; HIV risk factors affecting pregnancy)
- Educate patient:
  - About medication side effects and what to do should they occur.
    ▪ <u>Common</u>: diarrhea, dizziness, nausea, headache, fatigue, abnormal dreams, sleep problems, rash, depression.
      • Continue medication
      • Contact care team to discuss symptoms
      • Schedule an evaluation within one week of contact with care team
    ▪ <u>Serious</u>: lactic acidosis (weakness, unusual pain, trouble breathing, nausea, vomiting, fast heartbeat, feeling cold/dizzy/lightheaded), liver problems (jaundice, biliuria, light colored stools, lack of appetite, nausea, abdominal pain).
      • Discontinue medication
      • Contact care team to discuss symptoms that day
      • Schedule an evaluation within 24 hours of contact with care team
  - About acute HIV infection symptoms (fever, malaise, myalgia, rash, headache, night sweats, sore throat, lymphadenopathy), and what to do if these should occur.

##### f) Monitoring During Treatment

While on PrEP, the patient should be evaluated by the nurse one month after PrEP initiation and every 3 months thereafter. These quarterly visits shall include:

- Confirmation of HIV-negative status. This will entail obtaining a 4th generation HIV Antibody/Antigen test (and an HIV VL if the patient is symptomatic for acute HIV infection).
- Evaluation of PrEP medication adherence.
- Assessment of risk behaviors and provision of risk-reduction counseling and condoms.
- Assessment of STI symptoms and, if present, testing and treatment for STIs, as needed (at least every six months).
- Three months after initiation, then every six months while on PrEP medication, BUN and serum creatinine should be checked.
- In women of reproductive age and trans-males, a pregnancy test and, if pregnant, discussion of continued use of PrEP with patient.

## Footnotes

1 For the purposes of this paper, the term “women” will refer to people assigned female at birth

